# Alpha7 nicotinic acetylcholine receptor agonist PHA 568487 dampens inflammation in PBMCs from patients with newly discovered coronary artery disease

**DOI:** 10.1101/2024.03.16.24304336

**Authors:** Filip Mjörnstedt, Rebecka Wilhelmsson, Marcus Ulleryd, Maria Hammarlund, Göran Bergström, Anders Gummesson, Maria E Johansson

**Affiliations:** Department of Physiology, Institute of Neuroscience and Physiology, Gothenburg University, Gothenburg, Sweden; Department of Molecular and Clinical Medicine, Institute of Medicine, Sahlgrenska Academy, Gothenburg University, Gothenburg, Sweden; Department of Clinical Physiology, Sahlgrenska University Hospital, Västra Götalandsregionen, Gothenburg, Sweden; Department of Clinical Genetics and Genomics, Sahlgrenska University Hospital, Västra Götalandsregionen, Gothenburg, Sweden

**Keywords:** coronary artery disease, atherosclerosis, inflammation, alpha7 nicotinic acetylcholine receptor

## Abstract

**Background:** The alpha7 nicotinic acetylcholine receptor (α7nAChR) controls inflammation in experimental models. The α7nAChR is expressed in human peripheral blood mononuclear cells (PBMCs) as well as in human atherosclerotic plaques, yet, its role in regulating inflammation in patients with cardiovascular disease remains unknown. In this study we aim to investigate whether stimulation of the α7nAChR can dampen the immune response in patients with newly discovered coronary artery disease (CAD).

**Methods:** Human peripheral blood mononuclear cells (PBMCs) extracted from patients with verified CAD (n=38) and control participants with healthy vessels (n=38) were challenged in vitro with lipopolysaccharide (LPS) in combination with α7nAChR agonist PHA 568487. Supernatants were analyzed for cytokines using multiplex immunoassay. The CAD group was re-examined after 6 months.

**Results:** α7nAChR stimulation decreased TNF-α in all groups, in control participants and in CAD patients, both at the first visit as well as the follow-up visit after 6 months. The most pronounced effect of α7nAChR stimulation was seen in CAD patients at their first visit, where 12 of 17 cytokines were decreased (TNF-α, IL-1β, IL-2, IL-4, IL-5, IL-7, IL-10, IL-17A, GM-CSF, MCP-1, MIP-1β and IL12(p70)).

**Conclusions:** **S**timulation of α7nAChR dampens the inflammatory response in human PBMCs. This suggests that the anti-inflammatory properties of the α7nAChR may have a role in treating CAD.

## Introduction

Signalling via the alpha7 nicotinic acetylcholine receptor (α7nAChR) can regulate inflammation. Following activation of the cholinergic anti-inflammatory pathway, or via direct stimulation with agonists, the α7nAChR can suppress the release of pro-inflammatory cytokines from immune cells^1^. In humans, the α7nAChR is expressed both in peripheral blood mononuclear cells (PBMCs)^2^ and in atherosclerotic plaques^3^. Stimulation of the α7nAChR reduces the inflammatory response in whole blood from healthy donors^4^. However, it is unclear whether stimulation of α7nAChR can affect the inflammatory response in patients with coronary artery disease (CAD). This study aims to investigate the impact of α7nAChR stimulation, using the selective agonist PHA548687, on the immune response in PBMCs from both healthy individuals and patients with newly discovered CAD.

## Methods

A total of 38 patients with extensive coronary atherosclerosis (CAD) and 38 participants with no atherosclerosis (Controls) were recruited from The Swedish CardioPulmonary BioImage Study (SCAPIS)^5^. The study protocol was approved by the ethical review board in Gothenburg, Sweden and informed consent was obtained from all participants. Inclusion-criteria for CAD: three or more of the central segments 5, 6, 7, 1 or 11 having atherosclerosis or one of these segments with significant stenosis (>50%). When calcium blooming was present, the level of stenosis was set to 1-49%. Inclusion-criteria for controls: no signs of coronary atherosclerosis (negative CCTA and CAC scan). Exclusion criteria: History of myocardial infarction and/or angina pectoris or any symptoms of angina pectoris. Pharmacological treatment with statins within last 12 months. Any major surgical procedure or trauma within 4 weeks of the first study visit.

During the first study visit (CAD1), the CAD group met with a study physician, received lifestyle advice, and were recommended lipid-lowering statins to reduce the risk of future cardiovascular events. The CAD group was invited to a second visit (CAD2) six months later while the control group only attended one visit. Blood samples were drawn during the visits for biochemical analysis and PBMC extraction.

For the in vitro stimulations, PBMCs were seeded at a concentration of 2×10^5^ cells/well in triplicates and stimulated with LPS (8.33 ng/ml, List Biological Laboratories Inc., Campbell, USA) with/without α7nAChR agonist PHA568487 (83 μM) for 4 hours as previously described^2^. Supernatants were analyzed using the Bio-Plex Pro Human Cytokine Panel 17-plex assay (Bio Rad Laboratories, Inc. Hercules, USA) following the manufacturer’s protocol. Inclusion criteria for each participant: Following LPS stimulation, TNF response > 200 pg/ml and 10 times higher compared to unstimulated controls receiving PBS. PBMC viability after stimulation with PHA568487 was determined using alamarBlue Cell Viability Reagent (Invitrogen, Waltham, MA, USA) according to the manufacturer’s protocol. Two doses of PHA568487 (10μM or 100μM) were tested in human PBMCs obtained from healthy volunteers (blood bank Sahlgrenska University Hospital, n=4) and viability was measured after 24 h. No effect on viability was observed (data not shown).

Data were tested for normality with Shapiro Wilk’s test and statistical methods chosen accordingly. Population characteristics were analysed using one-way ANOVA followed by Sidak’s multiple comparison test. Cytokine data was analysed using one-way ANOVA mixed-effect analysis with Geisser-Greenhouse correction, followed by Holm-Sidak’s multiple comparison test. A significance level of p<0.05 was used. Differences in sample numbers were due to lack of sample material or lack of LPS response. The statistical analyses were conducted using GraphPad Prism software (La Jolla, USA) and IBM SPSS Statistics (NY, USA).

## Results

Statin treatment reduced cholesterol and LDL levels in CAD2 (Figure 1A). Among the three groups, only one cytokine, TNF, was significantly reduced (Figure 1B-D). In Controls the α7nAChR agonist PHA568487 reduced TNF levels while IL-6 levels were increased (Figure 1B). In CAD1, PHA568487 reduced the concentration of 12 out of the 17 cytokines analyzed (Figure 1C) whereas in CAD2, only TNF was reduced after PHA568487 treatment (Figure 1D). When dividing the CAD2 group into the two treatment categories, statin and lifestyle advice (n=20) versus lifestyle advice only (n=9), PHA568487 decreased the concentration of five cytokines in PBMCs from patients receiving lifestyle advice only (Fig. 1E). The LPS response was similar in Controls, CAD1 and CAD2. Likewise, there was no difference when comparing the LPS response of patients receiving lipid-lowering treatment and lifestyle advice to those receiving lifestyle advice only.

**Figure 1.**
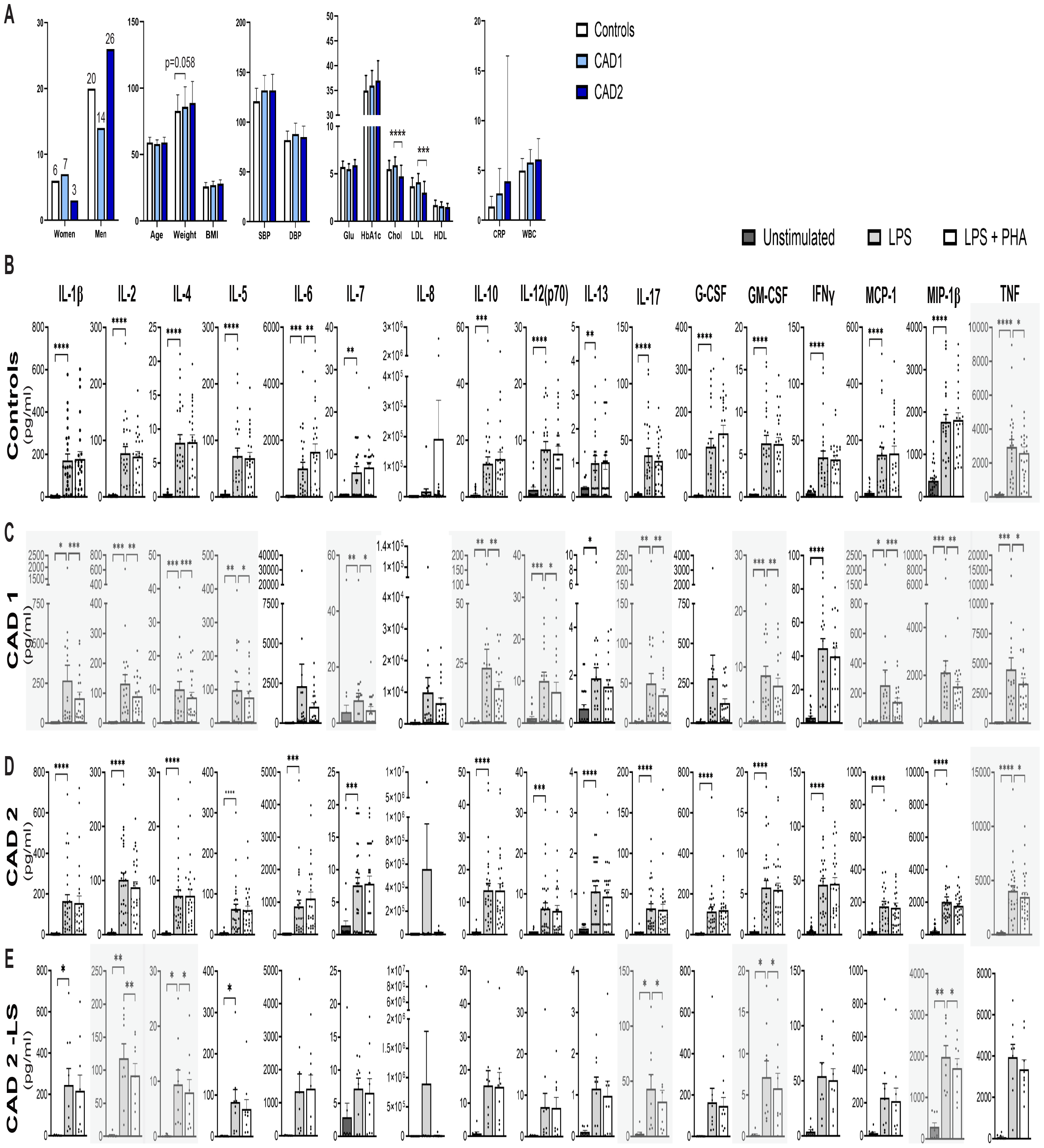
Stimulation of the alpha7 nicotinic acetylcholine receptor (a7nAChR) decreases the inflammatory response in PBMCs from patients with newly discovered coronary artery disease. PBMCs were challenged with LPS (8.33 ng/ml) and co-treated with α7nAChR agonist PHA-568487 (83 μM) for 4 hours. (A) Population characteristics. First panel from left; distribution of women and men included in the study, numbers above bars shows exact numbers of participants, second panel from left; age, weight (kg) and body mass index (BMI, kg/m^2^), third panel from left; systolic and diastolic blood pressure (SBP, DBP), fourth panel from left; glucose (Glu), HbA1c, LDL and HDL (all in mmol/L), fifth panel from left C-reactive protein (CRP, mg/L) and white blood cell (WBC, 10^9^/L). Data expressed as mean ± SD. (B-E) Multiplex analysis of cytokines IL-1β, IL-2, IL-4, IL-5, IL-6, IL-7, IL-8, IL-10, IL-12(p70), IL-13, IL-17A, G-CSF, GM-CSF, IFNγ, MCP-1, MIP-1β and TNF in (B) Controls (n=25-26), (C) CAD visit 1 (CAD1, n=19-21), (D) CAD visit 2 (CAD2, n=28-29) and (E) CAD visit 2 showing PBMC response from patients who only received life-style advice (CAD2-LS, n=9). Cytokine data expressed as mean ± SEM. Shaded areas highlights cytokines significantly decreased by α7nAChR agonist PHA568487. *p < 0.05, **p < 0.01, ***p < 0.001, ****p < 0.0001.

## Discussion

To the best of our knowledge, this is the first study to demonstrate a significant anti-inflammatory effect of α7nAChR stimulation on PBMCs from patients with newly discovered CAD. The study also confirms previous anti-inflammatory effects of α7nAChR stimulation in whole blood from healthy controls^4^. Interestingly, after six months of statin treatment, the anti-inflammatory effect of α7nAChR stimulation was reduced. Furthermore, patients receiving lifestyle advice had a greater anti-inflammatory effect of PHA56847 compared to patients receiving statins and lifestyle advice. These findings implicate that statins influence the inflammatory response. Indeed, statins not only reduce LDL levels in patients, but also inflammation by lowering the inflammatory biomarker hsCRP^6^. Furthermore, statins can suppress cytokine secretion from PBMCs; in hyperlipidemic patients, simvastatin treatment reduced secretion of IL-1β from PBMCs^7^. Intriguingly, the effect of α7nAChR stimulation is diminished after statin treatment. This lack of effect may be due to shared signaling pathways between the two, as both statins^8^ and α7nAChR agonists^9^ have been suggested to exert their anti-inflammatory effects by reducing the activity of the transcription factor nuclear factor-κB. Another possibility is that statins could alter the expression level of α7nAChR^9^. Surprisingly, IL-6 was increased in Controls, which may be explained by the fact that IL-6 is a pleiotropic cytokine that can both increase and reduce inflammation^10^. A limitation of the current study is that some participants were excluded due to low cell numbers from PBMC isolation or lack of LPS response, which prevented us from conducting a paired t-test analysis. Despite this limitation, the number of participants in each group was sufficient to identify significant differences.

In conclusion, stimulation of α7nAChR with PHA568487 reduces the inflammatory response of human PBMCs. This effect is more prominent in PBMCs from patients with newly discovered CAD, demonstrating the potential of α7nAChR’s anti-inflammatory properties in treating chronic inflammatory diseases such as CAD.

## Data availability

The participant-level datasets used for this report have been deposited with the Swedish National Data Service (https://snd.gu.se/, a data repository certified by Core Trust Seal). Due to patient consent and confidentiality agreements, the datasets can be made available for validation purposes by contacting. Data access will be evaluated according to Swedish legislation. Data access for research related questions in the program can be made available by contacting the corresponding author.

---

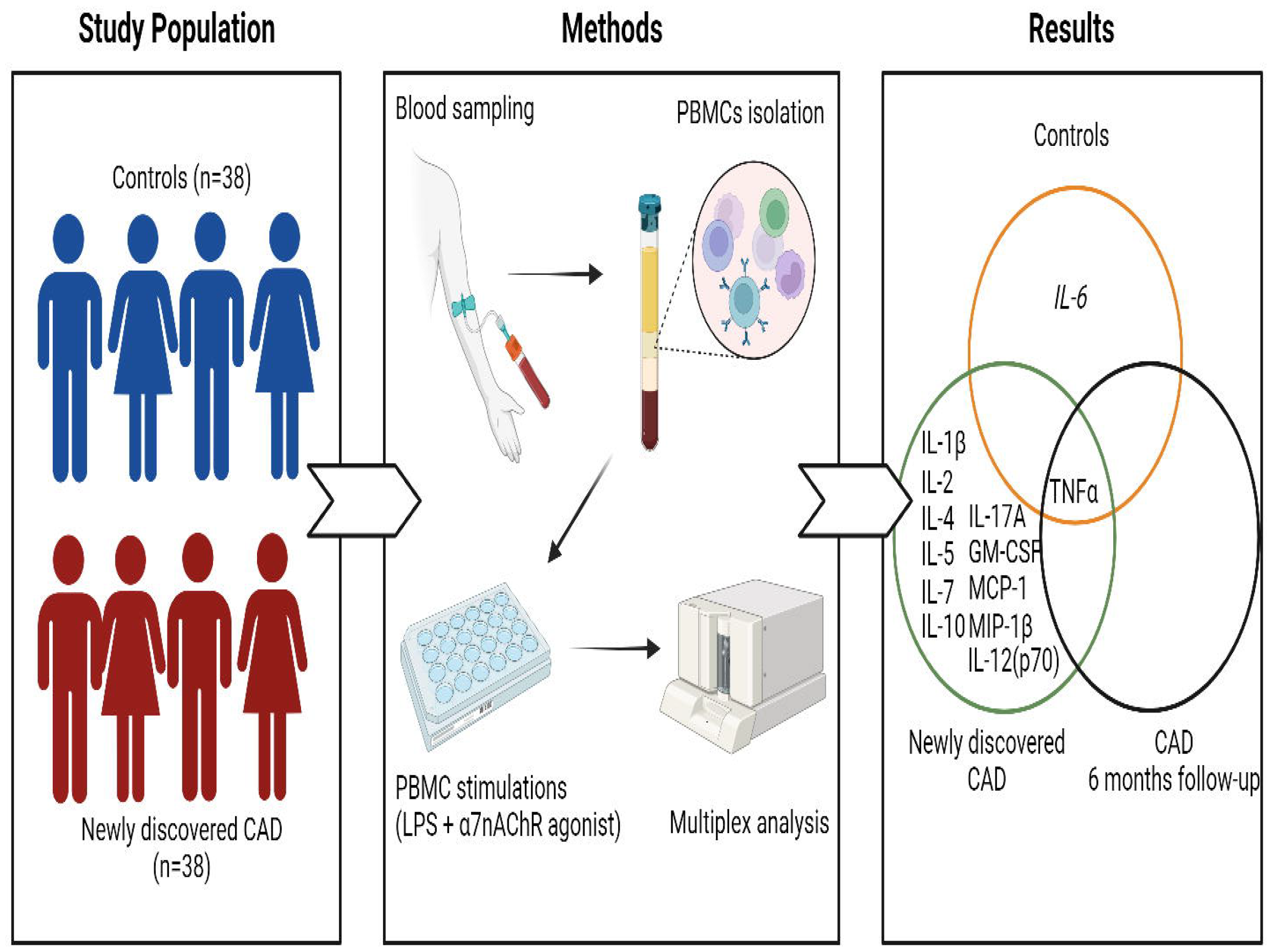

